# Carbapenem-resistant *Acinetobacter baumannii* at a hospital in Botswana: Detecting a protracted outbreak using whole genome sequencing

**DOI:** 10.1101/2023.07.10.23292487

**Authors:** Jonathan Strysko, Tefelo Thela, Andries Feder, Janet Thubuka, Tichaona Machiya, Jack Mkubwa, Kagiso Mochankana, Celda Tiroyakgosi, Kgomotso Kgomanyane, Tlhalefo Dudu Ntereke, Tshiamo Zankere, Kwana Lechiile, Teresia Gatonye, Chimwemwe Viola Tembo, Moses Vurayai, Naledi Mannathoko, Margaret Mokomane, Ahmed M Moustafa, David M Goldfarb, Melissa Richard-Greenblatt, Carolyn McGann, Susan E Coffin, Corrado Cancedda, Ebbing Lautenbach, Dineo Bogoshi, Anthony M Smith, Paul J Planet

## Abstract

Carbapenem-resistant *Acinetobacter baumannii* (CRAb) has emerged as a major and often fatal cause of bloodstream infections among hospitalized patients in low- and middle-income countries (LMICs). CRAb outbreaks are hypothesized to arise from reservoirs in the hospital environment, but outbreak investigations in LMICs are seldom able to incorporate whole genome sequencing (WGS) due to resource limitations. We performed WGS at the National Institute for Communicable Diseases (Johannesburg, South Africa) on stored *A. baumannii* isolates (n=43) collected during 2021–2022 from a 530-bed referral hospital in Gaborone, Botswana where CRAb infection incidence was noted to be rising. This included blood culture isolates from patients (aged 2 days – 69 years), and environmental isolates collected at the hospital’s 33-bed neonatal unit. Multilocus sequence typing (MLST), antimicrobial/biocide resistance gene identification, and phylogenetic analyses were performed using publicly accessible analysis pipelines. Single nucleotide polymorphism (SNP) matrices were used to assess clonal lineage. MLST revealed 79% of isolates were sequence type 1 (ST1), including all 19 healthcare-associated blood isolates and three out of five environmental isolates. Genes encoding for carbapenemases (*bla*_NDM-1_, *bla*_OXA-23_) and biocide resistance (*qacE*) were present in all 22 ST1 isolates. Phylogenetic analysis of the ST1 clade demonstrated spatial clustering by hospital unit. Nearly identical isolates spanned wide ranges in time (>1 year), suggesting ongoing transmission from environmental sources. One highly similar clade (average difference of 2.3 SNPs) contained all eight neonatal blood isolates and three environmental isolates from the neonatal unit. These results were critical in identifying environmental reservoirs (e.g. sinks) and developing remediation strategies. Using a phylogenetically informed approach, we also identified diagnostic genes useful for future tracking of outbreak clones without the need for WGS. This work highlights the power of South-South and South-North partnerships in building public health laboratory capacity in LMICs to detect and contain the spread of antimicrobial resistance.

## Introduction

Antimicrobial resistance (AMR) disproportionately affects low- and middle-income countries (LMICs), with sub-Saharan Africa bearing the highest burden.(1) Carbapenem antibiotics are considered last-line agents for infections with multidrug-resistant Gram-negative organisms, but the prevalence of carbapenem resistance is now rapidly increasing globally.(2) In 2019, carbapenem-resistant *Acinetobacter baumannii* (CRAb) was estimated to be the fourth leading pathogen responsible for AMR-attributable mortality and the World Health Organisation has named it a priority-one pathogen for research and development of new antibiotics.(1, 3)CRAb has been closely linked with healthcare settings, owing to its unique ability to evade killing by disinfectants and antimicrobials and to withstand desiccation on wet and dry surfaces.(4) Intensive care units (ICU) in resource-limited settings can be epicenters of environmental CRAb proliferation due to limited infection prevention and control (IPC) capacity to interrupt transmission among patients who may be immunocompromised and have indwelling medical devices.(5)

Surveillance data from LMICs demonstrate a pattern of CRAb disproportionately affecting neonates and infants. According to 2017-2019 laboratory-based sentinel surveillance data from South Africa, infants made up the highest proportion of *A. baumannii* infections, the majority of which were carbapenem-resistant.(6) A recent review of neonatal sepsis in seven LMICs in sub-Saharan Africa and south Asia reported that *A. baumannii* was among the top five etiologies in all countries surveyed.(7) Similarly, CRAb is now one of the top three causes of neonatal sepsis among hospitalized neonates in India and South Africa.(8-10) At the tertiary referral hospital in Botswana where this study was carried out, bloodstream infection surveillance records indicated that, between 2012 and 2021, the proportion of *A. baumannii*-attributable neonatal sepsis rose from 1% to 16% and was associated with a case fatality rate of 56%. (11, 12).

Molecular analyses have been used to detect protracted outbreaks of CRAb in settings where it has become hyperendemic and have helped to shed light on transmission pathways.(13-16) Whole genome sequencing (WGS), once considered cost-prohibitive, is proving to be a promising tool for outbreak detection in LMICs when traditional epidemiologic approaches fall short. WGS enables a range of analytic tools to characterize strain relatedness and identify relevant gene mutations encoding for antimicrobial and biocide resistance. Genes encoding for carbapenemases (β-lactamases (*bla*) including oxallase (OXA), and metallo-β-lactamases like *bla*_NDM-1_) are increasingly common among hospital CRAb isolates; the *bla*_OXA–23_ gene is the most predominant gene encoding for carbapenemase in CRAb, and is carried on various mobile elements including transposons, plasmids, and resistance islands.(17) Resistance to commonly-used biocides like quaternary ammonium compounds (qAC) may contribute to environmental persistence CRAb.(18)

In this study, we aimed to conduct WGS analysis on clinical and environmental *A. baumannii* isolates collected from a tertiary referral hospital in Botswana where anecdotal reports of CRAb infections were noted to be rising. We also present a set of diagnostic genes that can be used to track the clones identified here without the need for WGS.

## Materials and Methods

We performed WGS on all viable stored *A. baumannii* isolates collected from 1 March 2021 to 31 August 2022 at a 530-bed public tertiary referral hospital in Botswana with an 8-bed ICU and a 33-bed neonatal intensive care unit (NICU). During this period, *A. baumannii* isolate storage was coordinated by a laboratory-based surveillance program which had undergone ethical review and approval by the Health Research and Development Committee at Botswana’s Ministry of Health and Wellness and the Institutional Review Boards (IRB) at University of Botswana, University of Pennsylvania, and the healthcare facility where this study was carried. Isolates were abstracted on 1 September 2022 (authors did not have access to identifiable data) and were submitted to the National Institute for Communicable Diseases (Johannesburg, South Africa), for WGS. We obtained a waiver of written patient consent by the IRBs due to this being a retrospective abstraction of surveillance data. The entire process, from sample collection to genomic analyses, is illustrated in Figure 1.

**Figure 1.**
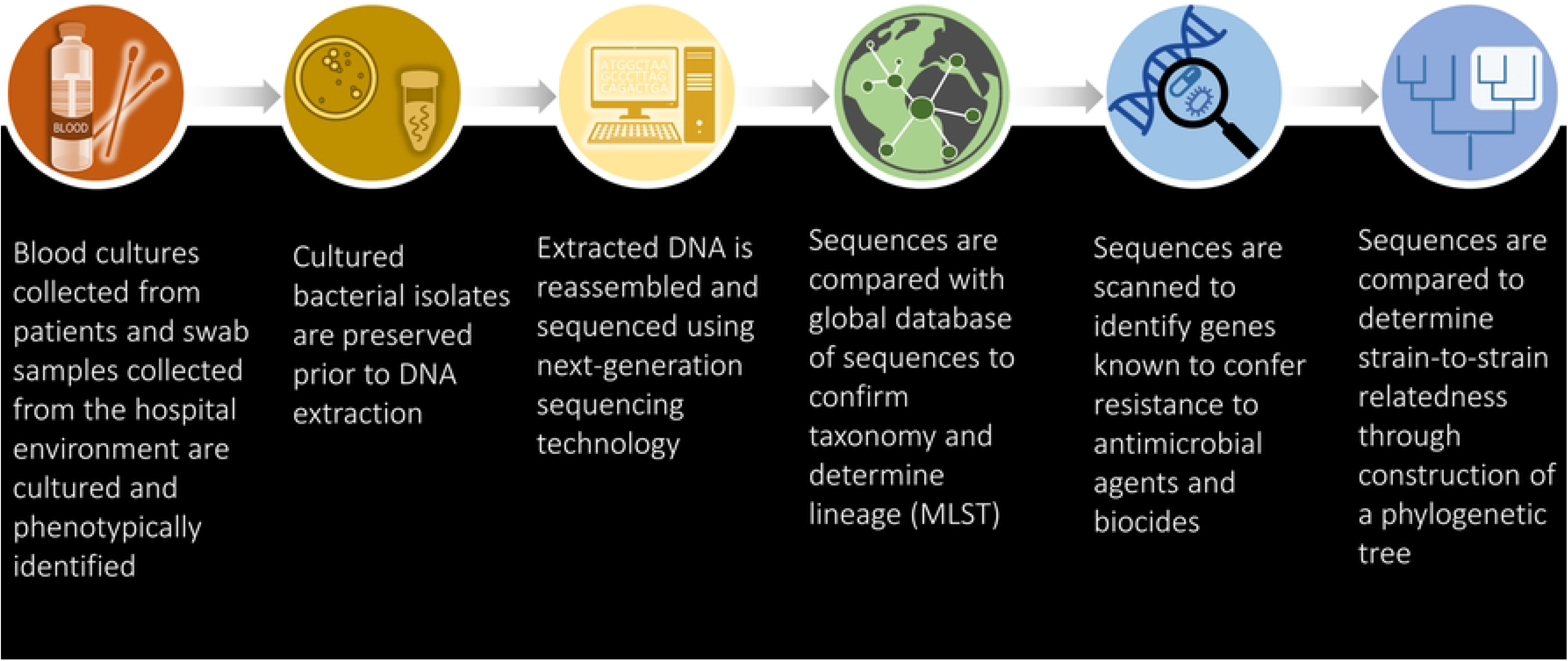
Sequence of events: From specimen collection to genomic analysis.

### Clinical isolates

Blood cultures were collected by hospital staff following appropriate skin antisepsis from patients when clinically indicated. These were then incubated at the hospital microbiology laboratory using an automated system (BACT/ALERT®, BioMérieux). Isolate identification was performed manually using Gram stain and colony morphology, followed by sub-culturing and analytical profile index strip testing or a combination of motility, indole, hydrogen sulphide, citrate, urease, and lysine testing. Blood cultures were classified as healthcare-associated if they were collected more than 72 hours after hospitalization or, in the case of neonates born at and never having left this healthcare facility, after the first day of life. All available stored blood culture isolates previously identified as *A. baumannii* were sub-cultured prior to submission for sequencing.

### Environmental isolates

Isolates presumptively identified as *Acinetobacter* spp. from banked environmental specimens collected as part of point prevalence surveys in this hospital’s NICU from April to June, 2021 were also revived and submitted for sequencing. Sites sampled included high-touch surfaces, water, equipment, and hands of caregivers and healthcare workers. Samples were collected using sterile nylon flocked swabs from sites without regard to epidemiologic links to patients with infection. Samples were transported in sterile water and individually inoculated onto chromogenic media within 24 hours for the selection and differentiation of extended-spectrum β-lactamase-producing organisms (CHROMagar™ ESBL, Paris, France). Colonies were identified using visual inspection of chromogenic media only.

### Whole Genome Sequencing

Following initial phenotypic identification by the hospital’s clinical microbiology laboratories, all isolates were stored at -80° C until shipment of isolates to NICD laboratories. At NICD, genomic DNA was extracted from bacteria using an Invitrogen PureLink Microbiome DNA Purification Kit (Invitrogen, USA). WGS (Illumina NextSeq, USA) was performed and DNA libraries were prepared using a Nextera DNA Flex Library Preparation Kit (Illumina), followed by 2 × 150 base pair paired-end sequencing runs with ∼80 times coverage. All WGS methodologies were performed using the protocols and standard operating procedures as prescribed by the manufacturers of the kits and equipment.

### Genome and Phylogenetic Analysis

Taxonomic assignment, MLST, antimicrobial resistance gene identification, and phylogenetic analyses were performed using publicly accessible analysis pipelines at NICD and at the University of Pennsylvania / Children’s Hospital of Philadelphia Microbiome Center. A maximum likelihood tree was built using the Cladebreaker pipeline and included the genomes from our collection plus 16 additional assembled genomes available on GenBank,(19) chosen using the topgenome (-t) feature of WhatsGNU with 3 top genomes specified.(20) A WhatsGNU database for *A. baumannii* was constructed from 14,627 publicly available genomes downloaded using National Center for Biotechnology Information genome download.(21) Bootstrap values, referring to the number of times the phylogenetic branching point is re-observed after repeated sampling of the data with replacement, were assigned after 100 repetitions of sampling from nonparametric pseudoreplicates.(22) SNP matrices were used to assess clonal lineage. There are no universal thresholds for determining whether any two CRAb strains are part of the same outbreak, but SNP differences of less than three have been used to conservatively define isolates as unequivocally part of an intra-hospital outbreak. (23) Genome-wide association studies were conducted using a script which performs case/control allele scoring based on WhatsGNU reports.(20) A full list of analysis pipelines used in genomic analysis is included in S Table 1.

## Results

Of 54 blood culture isolates identified as *A. baumannii* during the surveillance period at this hospital, a total of 23 isolates were available for sequencing. Cultures had been taken from patients aged 2 days to 69 years, and most (82%; 19/23) were healthcare-associated. All 23 blood culture isolates were confirmed to be *A. baumannii* by WGS. Out of 20 environmental isolates identified as *A. baumannii* using chromogenic media, only 5 were confirmed as *A. baumaannii*. The remaining environmental isolates were identified as other *Acinetobacter* spp. (n=3), *Serratia* spp. (n=6), *Pseudomonas* spp. (n=3), *Exiguobacterium* spp. (n=1), *Microbacterium* spp. (n=1), and an unclassified organism.

### Sequence typing and characterization

MLST analysis revealed 79% (22/28) of *A. baumannii* isolates to be sequence type 1 (ST1), including all 19 healthcare-associated blood isolates and 60% (3/5) of the environmental isolates, and phylogenetic analysis confirmed the monophyly of these genomes (S Figure 1). None of the 4 community-associated blood isolates were identified as ST1 and were unrelated to each other (ST976, ST193, and two unidentified MLST). Genes encoding for carbapenemases (*bla*_NDM-1_, *bla*_OXA-23_) and biocide resistance (*qacE*) were present in all 22 ST1 isolates; mutations associated with colistin resistance were not identified.

### Phylogenetic analysis

Phylogenetic analysis of the ST1 clade demonstrated spatial clustering by hospital unit. Very closely related isolates spanned wide ranges in time (>1 year), suggesting ongoing transmission from environmental sources (Figure 2). A neonatal clade (average difference 2.3, range 0-5 SNPs) containing all eight neonatal blood isolates were closely associated with three environmental isolates from the neonatal unit: a sink drain, bed rail, and a healthcare worker’s hand. Three isolates from one adult patient, from 3 separate blood cultures sent on the same day, were also included in this clade, however there was no clear epidemiologic link identified among the neonatal patients. The tree also suggested the existence of a local clade that contains both the neonatal clade as well as other isolates from the same hospital (average difference 13.8, range 0-30 SNPs).

**Figure 2.**
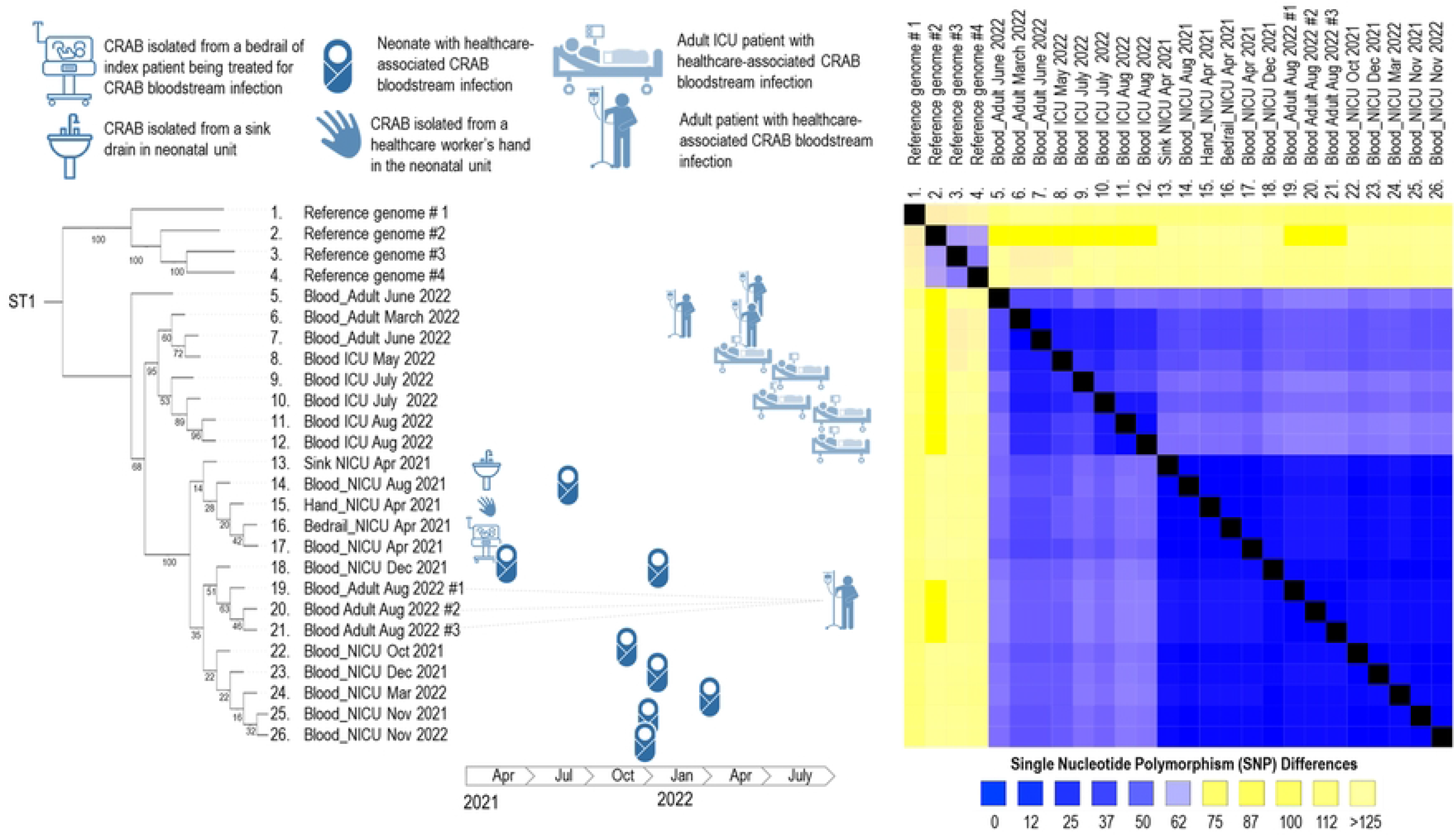
**Phylogenetic tree (left) with branch points labelled with bootstrap values (0-100) and single nucleotide polymorphism differences (right) of sequence type 1 (ST1) clinical and environmental carbapenem-resistant *Acinetobacter baumannii* (CRAb) isolates collected from April 2021 – August 2022**

### Diagnostic Alleles

Using the program WhatsGNU,(20) we interrogated our whole genome datasets for diagnostic protein alleles that defined each of the clades in our tree. We required each allele to have 100% specificity and 100% sensitivity for the genomes in each clade, and a GNU score of 0 indicating that this allele is never seen in the *Acinetobacter baumanii* whole genome database. The neonatal clade was characterized by 11 protein alleles with various predicted functions, and the local ST1 clade that includes the neonatal clade was characterized by 10 protein alleles most of which had no predicted function (S2 Figure). Nucleotide and amino acid sequences for each of these genes is included as supplementary data (S3 Appendix).

## Discussion

We detected a protracted hospital CRAb outbreak with transmission spanning at least 18 months using WGS. All healthcare-associated strains shared the same clonal lineage and many occurred in newborns who had never left the hospital, suggesting that ongoing transmission was facilitated by the presence of reservoirs in the hospital environment. Although only five environmental *A. baumannii* isolates were available for analysis, three shared the same clonal, extremely similar lineage as the healthcare-associated isolates and suggested that sink drains were likely reservoirs and hands of healthcare workers may have served as transmission vehicles.

Although the isolates sequenced generally clustered spatially by ward, their shared clonal lineage (ST1) and the case of an adult patient closely related to the “neonatal” clade raises questions around shared exposures across wards and reservoirs yet to be identified. These exposures could include: healthcare workers cross-covering wards, shared cleaning equipment (e.g. mops and buckets), or shared plumbing. Of note, no *A. baumannii* was isolated from fresh tap water samples obtained from water taps serving the NICU sinks during the environmental point prevalence surveys from which this analysis drew its environmental isolates.(24) This finding lends credence to the hypothesis that contamination arising from the sink is likely due to retrograde splashing of water from the sink drain onto either a patient or healthcare worker rather than from intrinsically contaminated municipal tap water; backsplash is a likely source of contamination of washbasins, tap handles, hands, and environment immediately surrounding a sink. Our study is amongst the first in an LMIC setting to utilize temporal sequencing of clinical and sink drainage isolates to demonstrate that the sink drains are a likely reservoir.(25)

Sinks, although important for hand hygiene, are often used for other purposes, and can harbor robust biofilms and serve as a reservoir for multidrug-resistant organisms.(26-28) These difficult-to-dislodge biofilms necessitate reservoir remediation strategies, especially for hospitals in resource-limited settings. Some evidence supports radical measures to eliminate water-related reservoirs including “water-free patient care” (i.e. complete removal of sinks from patient care areas).(29) However, these interventions have not been well-tested in high-resource settings, and their feasibility in resource-limited settings is unclear; sink re-design and remediation strategies (e.g. plumbing trap stop valves, boiling water, splashguards) may be more easily implemented in these settings.(30, 31)

Aside from reservoir remediation, there is a general need for rapid adaptation of IPC protocols appropriate for resource-limited settings. Use of invasive medical devices in LMICs has outpaced the IPC measures needed to maintain and safely reprocess them. Additionally, the global healthcare worker shortage has resulted in too few staff to implement IPC measures.(32) Multimodal IPC bundles are needed which deploy a combination of optimization of hand hygiene, colonization-based patient screening, environmental sampling, and enhanced terminal and non-terminal cleaning and disinfection.(33) Novel strategies to task shift some cleaning and disinfection activities to caregivers may also be worth consideration.

This study was conducted during the height of the COVID-19 pandemic and the extent to which the rise in CRAb incidence at this facility was related to the pandemic is unclear. Several regions in both high-income settings and LMICs reported an increase in CRAb incidence in 2020-2021, presumably as a result of the surge in ICU admissions during the COVID-19 pandemic.(34-36) Of note, the environmental point prevalence surveys referenced to in this study were conducted early in 2021 and, over the course of the six-month study, a temporal increase in *Acinetobacter* spp. recovery was observed, corresponding with both an increase in clinical CRAb infection and COVID-19 incidence hospital-wide.(24) The high prevalence of the *qacE* gene among CRAb isolates in this analysis also raise questions about the unintended consequences caused by the selective pressure of excess biocide use, particularly during the COVID-19 pandemic.(37) The lack of strain diversity over an extended period may suggest that *Acinetobacter* spp. is capable of persisting in the hospital environmental reservoirs at no fitness cost despite competition from other organisms or use of disinfectants.

This analysis highlights the power of WGS to solve complex public health problems in LMICs bearing the highest burden of AMR. As WGS technologies become cheaper and open-source analytic tools more accessible, broad support for the adaptation of WGS in public health laboratories in LMICs is needed.(38) South-South and North-South collaborations like these will be pivotal in supporting technical development and in ensuring sustainable implementation of WGS into national action plans to combat AMR.

Even as LMICs welcome WGS technology into the public health laboratory ecosystem, alternative, low-cost strategies are needed for molecular tracking in the midst of outbreaks like these. Identification of diagnostic alleles for clade typing described here, is an emerging field promising a low-cost method of tracking outbreak strains.(39) There are two major benefits to this diagnostic approach. First, new isolates can be screened for inclusion in each clade without full-scale phylogenetic analysis. Second, amplification of these genes and sequencing by standard Sanger sequencing can be done at a fraction of the cost of WGS. PCR primers could also be designed to amplify these genes directly from clinical samples without the need for culture. It may also be the case that the alleles detected here may have biological significance that would inform future studies about the fitness and biological properties of outbreak strains.

### Limitations

This study was limited by the small number of environmental *A. baumannii* isolates available for sequencing. Identifying genetically linked isolates from patient and environmental samples does not prove the sampled site is the definitive point-source. Because this clonal type was recovered from multiple environmental sites, it is likely that there may have been additional reservoirs and vehicles not identified because of the limited nature of the environmental sampling, thus obscuring the full spectrum of transmission. Future investigations would incorporate more samples and would prioritize sites epidemiologically linked to patients with confirmed infection. Additionally, the use of chromogenic media to screen for *A. baumannii* in environmental samples had low specificity; future studies would take advantage of media designed specifically for *A. baumannii* recovery, as well as automated identification of environmental isolates prior to sequencing.

## Conclusions

This report demonstrates the power of WGS to detect outbreaks when traditional epidemiologic approaches fall short. Additionally, our work makes a case for public health laboratories in LMICs to be equipped with sequencing technologies so WGS can be leveraged in the regions bearing the highest burden of AMR. South-South and North-South partnerships, like the ones described in this manuscript, are critical in building public health laboratory infrastructure to detect and contain health threats caused by AMR. This work also highlights the importance of environmental sampling to identify and remediate reservoirs (e.g. sink drains) and vehicles (e.g. hands, equipment) within the healthcare environment and is a call-to-action for broader support for IPC teams in resource-limited settings.

## Data Availability

The authors confirm that the data supporting the findings of this study are available within the article and its supplementary materials.

## Acknowledgments

Whole-genome sequencing of bacterial isolates was made possible by the SEQAFRICA project supported by the Department of Health and Social Care’s Fleming Fund using UK aid. The views expressed in this publication are those of the authors and not necessarily those of the UK Department of Health and Social Care or its Management Agent, Mott MacDonald.

